# The MHCII Immune Activation Score predicts risk of recurrence and benefit of taxanes in Basal-like and HER2-enriched breast cancer

**DOI:** 10.64898/2026.06.24.26356102

**Authors:** Philip S. Bernard, Bingshu E. Chen, Dongxia Gao, Lois E. Shepherd, Torsten O. Nielsen, Katherine E. Varley

**Affiliations:** Department of Pathology, Huntsman Cancer Institute, University of Utah, Salt Lake City, UT, USA; Canadian Cancer Trials Group, Queen’s University, Kingston, ON, Canada; Department of Pathology and Laboratory Medicine, BC Cancer and University of British Columbia, Vancouver, Canada; Department of Oncological Sciences, Huntsman Cancer Institute, University of Utah, Salt Lake City, UT, USA

## Abstract

**Purpose:** There are no clinically validated biomarkers to assess recurrence risk and guide treatment de-escalation in Basal-like and HER2-enriched breast cancer. Taxane-based chemotherapy remains a cornerstone of treatment despite significant toxicity. We evaluated the prognostic and predictive utility of the MHCII Immune Activation Score (IA Score) in these subtypes.

**Experimental Design:** We retrospectively analyzed Basal-like and HER2-enriched breast cancers from the NCIC CTG MA.21 trial, which randomized patients with node-positive or high-risk node-negative disease to adjuvant chemotherapy with or without taxanes. MA.21 predated immune checkpoint inhibitors and routine HER2-targeted therapy. Subtype was previously assigned by PAM50. The 36-gene MHCII-IA assay used RNA from formalin-fixed, paraffin-embedded tissue. Multivariable Cox and Kaplan-Meier analyses evaluated associations between IA Score, clinicopathologic variables, tumor-infiltrating lymphocytes (TILs), relapse-free survival (RFS), and taxane benefit.

**Results:** Among Basal-like (N=317) and HER2-enriched (N=155) tumors, higher IA Score was associated with improved RFS independent of lymph node status and provided stronger prognostic discrimination than TILs. Node-negative patients with high IA Score had excellent outcomes (8-year RFS >90%) versus those with low IA Score (8-year RFS <76%). In node-positive disease, high IA Score increased 8-year RFS by >10% relative to low IA Score. IA Score stratified taxane benefit: node-positive IA-low patients benefited, whereas IA-high tumors had favorable outcomes regardless of regimen.

**Conclusions:** MHCII Immune Activation Score is a prognostic and predictive biomarker in Basal-like and HER2-enriched breast cancer. High IA Score identified patients with excellent outcomes before pembrolizumab, trastuzumab, and taxane-based treatment escalation, providing a rationale for prospective risk-adapted de-escalation strategies.

## Introduction

Biomarker-driven precision medicine has become the cornerstone of modern breast cancer management. In clinical practice, immunohistochemical (IHC) evaluation of estrogen receptor (ER), progesterone receptor (PR), and human epidermal growth factor receptor 2 (HER2) remains the primary method for subtyping and therapeutic stratification, which approximates the main biological subtypes (Luminal A, Luminal B, HER2E and Basal-like) identified by PAM50 gene expression profiling^1^. For early-stage HR+/HER2- (Luminal) subtype, the last decade has seen a paradigm shift through the integration of multigene expression assays—such as Oncotype DX^2^, MammaPrint^3^, Breast Cancer Index^4^, Prosigna^5^, and EndoPredict^6^. Prospective clinical studies have demonstrated that these assays are prognostic and can aid clinical decision making for chemotherapy, extended endocrine therapy, and other adjuvant treatments ^7^. Their integration into clinical practice has led to significant improvements in patient care, including the safe de-escalation of chemotherapy and reduction of treatment-related toxicity in patients with favorable prognoses^8^.

In contrast, there are currently no widely adopted clinical biomarker tests to guide treatment de-escalation for patients with Basal-like Triple-Negative Breast Cancer (TNBC) or HER2-enriched (HER2E) HER2+ breast cancer. In these subtypes, chemotherapy remains the cornerstone of systemic treatment. Taxanes, such as paclitaxel, were incorporated into anthracycline-based regimens following clinical trials demonstrating modest but significant survival benefits^9-12^. However, taxane-based therapy is a primary driver of chemotherapy-induced peripheral neuropathy, a debilitating condition characterized by pain and numbness that affects up to 68% of patients one month post-treatment and persists as a permanent disability in many breast cancer survivors^13,14^. There is thus a critical, unmet need for biomarkers that can identify patients with an inherently favorable prognosis who may be safely spared the long-term morbidity of taxane-based regimens.

Host anti-tumor immunity has emerged as a key determinant of outcome in both Basal-like/TNBC and HER2E/HER2+ breast cancers^15,16^. Despite the well-recognized prognostic importance of tumor infiltrating lymphocytes (TILs) in early stage and metastatic ER-negative (TNBC and HER2+) breast cancers^16-18^, there are no recommended guidelines from CAP, ASCO, or NCCN for using TIL scoring alone for treatment planning. Stromal TIL scoring, which evaluates the global distribution of lymphocytes and plasma cells throughout the tumor’s connective tissue (continuous % of stromal area occupied, reported in 5-10% increments from H&E), has been shown to be an independent prognostic feature, even when considering molecular mechanistic biomarkers of immune response (e.g. PD-L1 IHC)^19^. The routine adoption of TIL assessment is hindered by inter-observer variability, lack of standardization in pathology reporting, no consensus on the optimal TIL threshold for clinical decision making, and the fact that it measures general immune cell abundance rather than functional activation^16,20^.

Clinical trials have relied on PD-L1 IHC for stratification and predicting response to immune checkpoint therapy^21,22^. For instance, KEYNOTE-355, a phase 3 clinical trial randomizing metastatic TNBC patients to chemotherapy plus pembrolizumab versus chemotherapy plus placebo, showed that pembrolizumab responders could be enriched using a PD-L1 immunohistochemistry combined positive score (CPS) ≥10 ^22^. The CPS is calculated by visually counting PD-L1-positive cancer cells, lymphocytes, and macrophages, divided by the total number of viable tumor cells, multiplied by 100. The reported agreement between CPS and TIL scoring in TNBC is relatively low (r∼0.4)^23^.

Tumor-cell expression of the Major Histocompatibility Complex Class II (MHCII) antigen presentation pathway represents a marker of functional immune activation^24-34^. The MHCII Immune Activation assay is a standardized multigene expression assay designed to measure this immune activation signature in formalin-fixed paraffin-embedded (FFPE) specimens^33^. This assay simultaneously measures MHCII antigen presentation (CIITA, NCOA1, CD74, CTSH, HLA genes), interferon-gamma cytokine activation (IFNG, IL7R, CD69), the presence of T cells (CD4, CD8A), and the PD-1/PD-L1 (PDCD1/CD274) checkpoint axis^33^. The MHCII-IA assay overcomes much of the variability in measurement and technical challenges of TIL assessment and IHC. Prior studies have demonstrated that an MHCII Immune Activation Score (IA Score) threshold can identify patients with very low risk of recurrence, achieving high specificity in independent validation cohorts^33^.

The NCIC CTG MA.21 trial (NCT00014222) provides a valuable framework to address this question. This international phase III study compared adjuvant cyclophosphamide, epirubicin, and fluorouracil (CEF) with regimens incorporating paclitaxel, including epirubicin and cyclophosphamide followed by paclitaxel (EC/T) and doxorubicin and cyclophosphamide followed by paclitaxel (AC/T)^35^. The primary endpoint was relapse-free survival (RFS). Initial analyses did not demonstrate a significant overall benefit of adding taxane therapy (EC/T vs. CEF)^35^. A subsequent study used the PAM50 assay^36^, the precursor of Prosigna, to classify tumor specimens from 1,094 MA.21 patients into intrinsic subtypes (Luminal A, Luminal B, HER2E, and Basal-like) and found that taxane benefit was restricted to non-Luminal subtypes^37^.

We hypothesize that high IA Scores identify a subset of Basal-like and HER2E tumors with intrinsically favorable prognosis and limited benefit from taxane-based therapy. In this study, we performed the MHCII Immune Activation assay on RNA remaining from the PAM50 subtyping of tumor specimens from the MA.21 trial. We also performed histological TIL assessment to compare the prognostic value of these immune biomarkers. The pre-specified primary objective of this study was to determine if high IA Scores are associated with significantly longer RFS in patients with Basal-like or HER2E tumors, regardless of chemotherapy regimen. The secondary objective was to determine if patients with high IA Scores experience comparably favorable outcomes in both the CEF and EC/T arms of the MA.21 trial and hence do not benefit from the addition of paclitaxel.

## Materials and Methods

### Study population

As described in previous publications, the NCIC CTG MA.21 trial (Registration ID: NCT00014222) was an international phase III trial that enrolled 2,104 patients between December 2000 and April 2005^35,37^. Following standard local control, patients were randomized to three adjuvant chemotherapy arms: doxorubicin, cyclophosphamide, and paclitaxel (AC/T); dose-intense cyclophosphamide, epirubicin, and fluorouracil (CEF); or dose-dense, dose-intense epirubicin, cyclophosphamide, and paclitaxel (EC/T). Patients were ≤60 years, with node-positive or high-risk node-negative disease, with median 8-year follow-up. The MA.21 primary end point was RFS defined as the time from randomization to the time of recurrence of the primary disease. Local or nodal recurrence and metastatic disease were considered a recurrence of the primary tumor. Patients who had contralateral breast cancer or a second primary malignancy or died from some cause other than disease were censored as relapse-free at time of death. Patients who had not relapsed were censored at longest follow-up.

### PAM50 subtype data

PAM50 subtyping data was collected for 1,094 MA.21 cases in a study that was published previously^37^. Patients enrolled in the MA.21 trial gave informed consent for acquisition of FFPE primary breast cancer tissue for correlative studies. Hematoxylin and eosin (H&E)-stained slides were reviewed by a pathologist blinded to identifiers or clinical outcome, and areas containing representative invasive breast carcinoma were marked on the H&E slides. Samples with no tumor present or insufficient tumor tissue area (<4 mm2) were excluded. Unstained serial tissue sections were macrodissected to remove the surrounding non-tumor tissue and total RNA was extracted from the unstained tumor tissue using the Roche High Pure RNA extraction kit (MilliporeSigma cat #: 04823125001). Samples that passed RNA extraction quality control criteria (RNA concentration ≥ 12.5 ng/µl and ratio of 260/280 = 1.70–2.50) were analyzed on the NanoString nCounter using probes specific for the PAM50 gene set. Raw data that passed quality metrics were sent to NanoString Technologies in a blinded fashion for PAM50 algorithm analysis, and subtype (luminal A, luminal B, HER2E, Basal-like) was assigned to each of the samples. RNA from patient tumors that were classified as Basal-like or HER2E were considered for this study.

### Approval for use of patient specimens

Approval for the use of archival specimens from the CCTG MA.21 Trial in this study was granted by the Breast Correlative Sciences Tumour Biology Working Group of the Canadian Cancer Trials Group, and the Institutional Review Board at the University of Utah (IRB # 157933).

TIL counting: Visual scoring of TILs was completed from full face section H&E images derived from the primary tumor excision specimens using the validated methodology described by the International TILs Working Group, performed by a single pathologist with extensive experience using this methodology (DG)^38,39^. This determines the percentage of stromal area within the bounds of invasive breast carcinoma that is occupied by mononuclear inflammatory cells^17^.

### MHCII Immune Activation Assay

Archival RNA specimens remaining from the PAM50 subtyping study^37^ were shipped from the Ontario Cancer Research Institute in Toronto, Ontario. The MHCII Immune Activation assay was applied to RNA specimens from primary patient tumors that were classified as Basal-like or HER2E by the previous PAM50 analysis^37^ and had sufficient RNA remaining (concentration >= 35.75 ng/uL and volume >= 8 uL).

The MHCII Immune Activation assay uses 36 custom gene-specific oligonucleotide probes (Integrated DNA Technologies) and Elements TagSets for measurement on the NanoString nCounter instrument^33^. The probe A oligos were HPLC purified and pooled at 5nM for each oligo in a Master Probe A Stock. The Probe B oligos were PAGE purified and pooled at 25nM for each oligo in a Master Probe B Stock. Before each NanoString nCounter run, 4 ul of each Master Probe Stock was diluted in 29 uL TE-Tween (10 mM Tris pH 8, 1 mM EDTA, 0.1% Tween-20) to make a Probe A Working Pool and a Probe B Working Pool. A Master Mix is then created by first adding 7 uL of Probe A Working Pool and 70 uL Hybridization Buffer to a NanoString nCounter Elements XT TagSet tube, then adding 7 uL of Probe B Working Pool. Hybridization reactions were then prepared for each RNA specimen by combining 7 uL of RNA at 35.7 ng/uL in water (total of 250 ng RNA) and 8 uL Master Mix for total volume of 15 uL. Probes were hybridized to the RNA at 67°C for 16 hours. Each run included 10 MA.21 RNA specimens, 1 positive control RNA, and 1 no-template ‘blank’ control. After hybridization, samples were transferred to the automated nCounter Prep Station for purification and immobilization onto the nCounter Master Kit cartridge. After sample preparation was complete, the cartridge was transferred to the nCounter Digital Analyzer for imaging and analysis. All samples were analyzed at 555 images per sample (FOV555).

Raw RCC files were imported into nSolver Analysis Software 4.0. The gene expression count values for each sample were normalized to correct for differences in background signal intensity across runs, and to correct for differences in RNA template quality and quantity between samples. A “no template” control sample was analyzed in each NanoString nCounter run and blank lane background subtraction was performed. Normalization across samples was then performed using the geometric mean of housekeeping genes included in the codeset (*ACTB, MRPL19, PSMC4, RPLP0, SF3A1*). The MHCII gene score for each sample was calculated using the geometric mean of the normalized counts for the following genes: *CD74, CIITA, CTSH, HLA-DMA, HLA-DMB, HLA-DPA1, HLA-DPB1, HLA-DPB2, HLA-DRB1*, and *NCOA1*. The TIL gene score for each sample was calculated using the geometric mean of the normalized counts for the following genes *ARHGAP9, CD274, CD3D, CD4, CD69, CD8A, IFNG, IL7R, PDCD1*. The IA Score for each sample was calculated as the geometric mean of the MHCII gene score and the TIL gene score.

### Statistical Analyses

The running of the MHCII Immune Activation assay and the calculation of TIL Score, MHCII Score, and IA Score were performed by individuals blinded to the patient survival data. Clinical and pathologic characteristics were summarized by intrinsic subtype and recurrence status using median (interquartile range) for continuous variables and counts (percentages) for categorical variables. Group comparisons were performed using the Wilcoxon rank-sum test for continuous variables and Fisher’s exact test for categorical variables.

Correlations between histologic tumor-infiltrating lymphocyte (TIL) scores and IA Scores were assessed using Spearman rank correlation. IA Scores were analyzed on the log10 scale for visualization where appropriate.

In accordance with the pre-specified statistical analysis plan, RFS was defined as the time from randomization to recurrence or censoring. Multivariable Cox proportional hazards models were used to evaluate associations between clinical variables (age, tumor stage, nodal status) and immune biomarkers (TILs or log10-transformed IA Score) within each intrinsic subtype. Results are reported as hazard ratios (HRs) with 95% confidence intervals (CIs).

To assess the incremental prognostic value of immune biomarkers, nested Cox models were compared using likelihood ratio tests, Akaike information criterion (AIC), and pseudo-R². Model discrimination was evaluated using the concordance index (C-index), and changes in C-index were calculated relative to the clinical model. The fraction of recurrences among patients in the lowest-risk quartile (based on model-derived linear predictors) was also computed.

Threshold-based analyses of recurrence risk were performed using a sliding-window approach across increasing values of TIL and IA Scores, stratified by nodal status and subtype. Generalized additive models were used to estimate smoothed recurrence probabilities and corresponding confidence intervals.

As described in the pre-specified statistical analysis plan, Kaplan–Meier methods were used to estimate RFS, with comparisons between groups performed using the log-rank test. Analyses were conducted using both a cohort-specific median IA cutpoint and a previously defined threshold (IA ≥ 2400). Additional subgroup analyses evaluated treatment effects (CEF vs. EC/T) stratified by IA Score and nodal status.

All statistical tests were two-sided, and p-values < 0.05 were considered statistically significant.

### Data Availability

The data, code, and compute environment used to produce the analyses and figures reported in this manuscript are available on Code Ocean https://codeocean.com/capsule/9802700/tree/v1.

## Results

### Cohort

The NCIC CTG MA.21 trial randomized 2,104 high-risk breast cancer patients to three arms: doxorubicin, cyclophosphamide, and paclitaxel (AC/T); dose-intense cyclophosphamide, epirubicin, and fluorouracil (CEF); or dose-dense, dose-intense epirubicin, cyclophosphamide, and paclitaxel (EC/T). PAM50 subtyping from a previous study^37^ and protocol-specific treatment data were available for 1,094 participants who were classified into Luminal A (n=299), Luminal B (n=256), HER2E (n=192), and Basal-like (n=347) groups. Figure 1 depicts downstream availability of biospecimens for the two subtypes analyzed in this study. Among HER2E tumors, 155 samples had sufficient remaining RNA for the MHCII IA assay, 155 passed NanoString QC, and 153 had H&E slides available for histologic TIL scoring. Among Basal-like tumors, 318 had sufficient remaining RNA, 317 passed NanoString QC, and 316 had H&E slides available for histologic TIL scoring.

**Figure 1.**
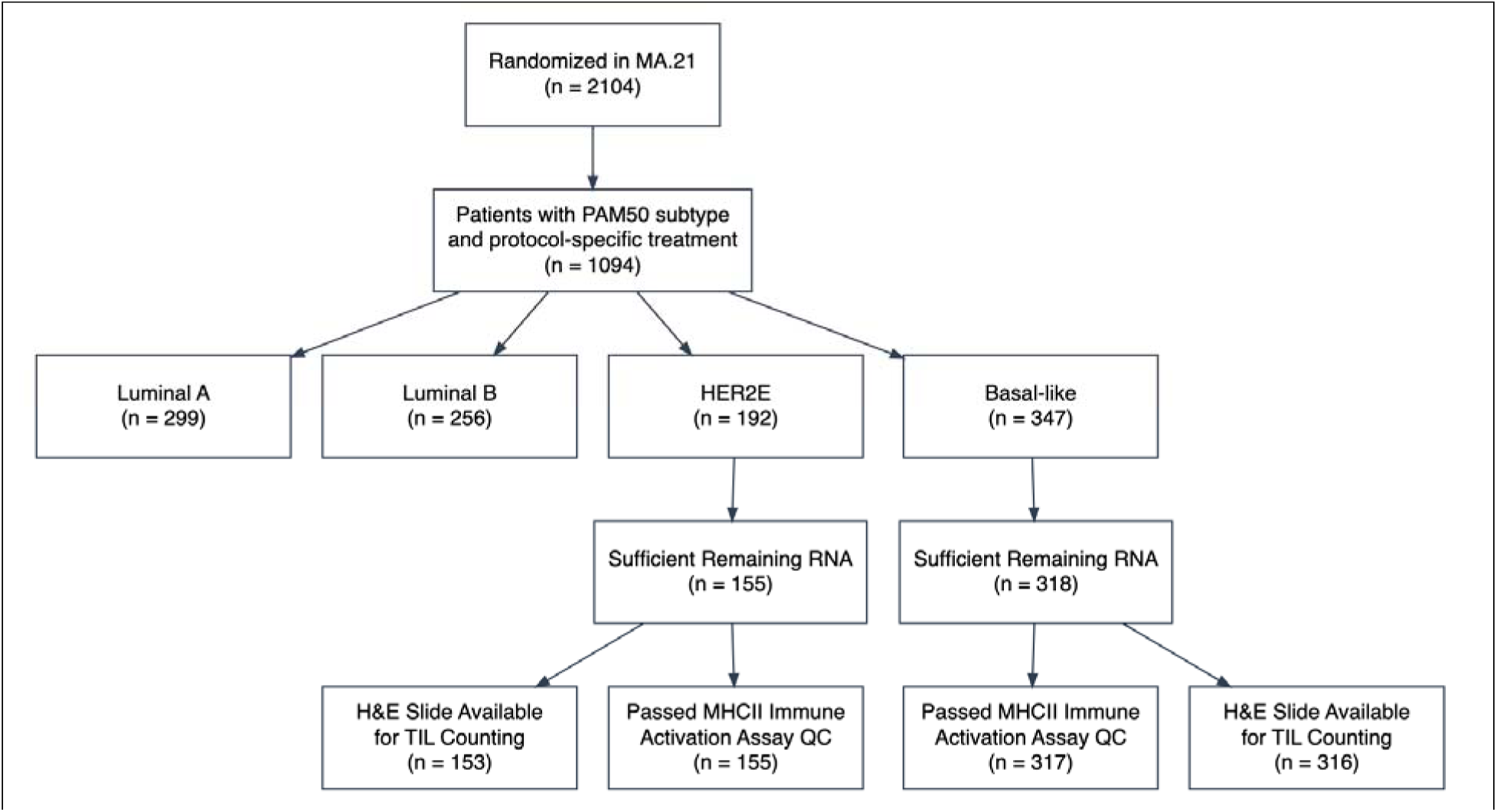
CONSORT diagram of MA.21 trial participants that had prior PAM50 subtyping, sufficient remaining RNA, MHCII Immune Activation assay data that passed NanoString Quality Control metrics and had H&E slides available for TIL counting for inclusion in analyses.

### Clinical covariates and immune biomarkers

A summary of clinical covariates and immune biomarker measurements is shown in Table 1 stratified by subtype and recurrence status. In both Basal-like and HER2E tumors, age and menopausal status did not differ significantly between patients who did and did not recur. Lymph node involvement was strongly associated with recurrence in both subtypes, with a higher proportion of node-positive disease among patients who experienced relapse (Basal-like, p = 0.003; HER2E, p = 0.005). In Basal-like tumors, both TIL and IA Scores were significantly higher in patients without recurrence (p = 0.007 and p = 0.005, respectively), whereas in HER2E tumors, only IA Score was significantly associated with RFS (p = 0.022).

**Table 1.**
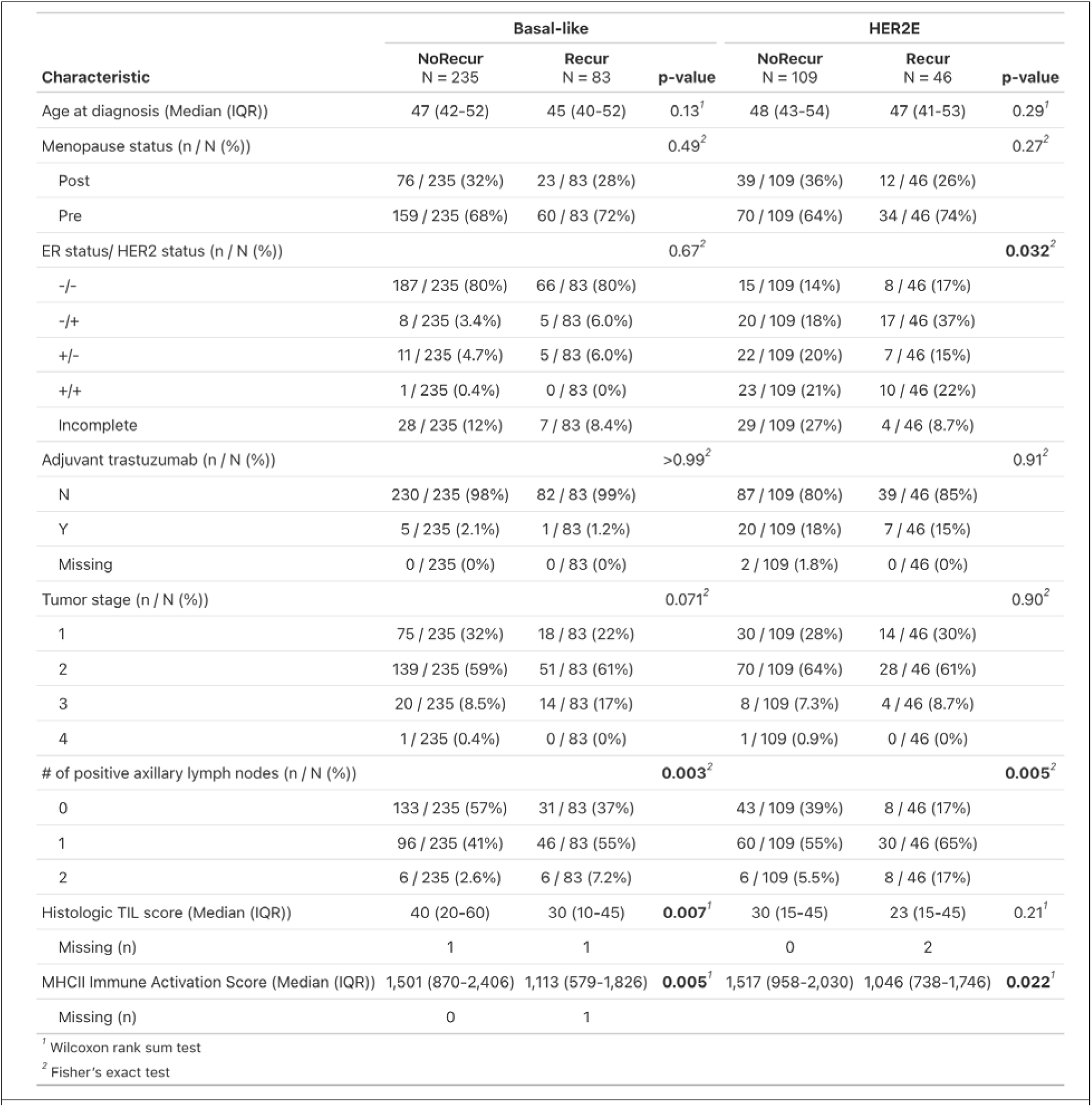
Patient and tumor characteristics by intrinsic subtype and recurrence status. Baseline clinical and immune covariates are shown for patients with Basal-like (left) and HER2E (right) tumors, stratified by relapse-free survival status (NoRecur vs Recur). Continuous variables are summarized as median (interquartile range), and categorical variables as n / N (%). P-values compare NoRecur vs Recur within each subtype using the Wilcoxon rank-sum test for continuous variables and Fisher’s exact test for categorical variables. Missing values are reported explicitly where present. Row-specific denominators reflect the number of evaluable patients for each variable.

| Characteristic | Basal-like |  |  | HER2E |  |  |
| --- | --- | --- | --- | --- | --- | --- |
|  | NoRecur<br>N = 235 | Recur<br>N = 83 | p-value | NoRecur<br>N = 109 | Recur<br>N = 46 | p-value |
| Age at diagnosis (Median (IQR)) | 47 (42-52) | 45 (40-52) | 0.13<sup>1</sup> | 48 (43-54) | 47 (41-53) | 0.29<sup>1</sup> |
| Menopause status (n / N (%)) |  |  | 0.49<sup>2</sup> |  |  | 0.27<sup>2</sup> |
| Post | 76 / 235 (32%) | 23 / 83 (28%) |  | 39 / 109 (36%) | 12 / 46 (26%) |  |
| Pre | 159 / 235 (68%) | 60 / 83 (72%) |  | 70 / 109 (64%) | 34 / 46 (74%) |  |
| ER status/ HER2 status (n / N (%)) |  |  | 0.67<sup>2</sup> |  |  | 0.032<sup>2</sup> |
| -/- | 187 / 235 (80%) | 66 / 83 (80%) |  | 15 / 109 (14%) | 8 / 46 (17%) |  |
| -/+ | 8 / 235 (3.4%) | 5 / 83 (6.0%) |  | 20 / 109 (18%) | 17 / 46 (37%) |  |
| +/- | 11 / 235 (4.7%) | 5 / 83 (6.0%) |  | 22 / 109 (20%) | 7 / 46 (15%) |  |
| +/+ | 1 / 235 (0.4%) | 0 / 83 (0%) |  | 23 / 109 (21%) | 10 / 46 (22%) |  |
| Incomplete | 28 / 235 (12%) | 7 / 83 (8.4%) |  | 29 / 109 (27%) | 4 / 46 (8.7%) |  |
| Adjuvant trastuzumab (n / N (%)) |  |  | >0.99<sup>2</sup> |  |  | 0.91<sup>2</sup> |
| N | 230 / 235 (98%) | 82 / 83 (99%) |  | 87 / 109 (80%) | 39 / 46 (85%) |  |
| Y | 5 / 235 (2.1%) | 1 / 83 (1.2%) |  | 20 / 109 (18%) | 7 / 46 (15%) |  |
| Missing | 0 / 235 (0%) | 0 / 83 (0%) |  | 2 / 109 (1.8%) | 0 / 46 (0%) |  |
| Tumor stage (n / N (%)) |  |  | 0.071<sup>2</sup> |  |  | 0.90<sup>2</sup> |
| 1 | 75 / 235 (32%) | 18 / 83 (22%) |  | 30 / 109 (28%) | 14 / 46 (30%) |  |
| 2 | 139 / 235 (59%) | 51 / 83 (61%) |  | 70 / 109 (64%) | 28 / 46 (61%) |  |
| 3 | 20 / 235 (8.5%) | 14 / 83 (17%) |  | 8 / 109 (7.3%) | 4 / 46 (8.7%) |  |
| 4 | 1 / 235 (0.4%) | 0 / 83 (0%) |  | 1 / 109 (0.9%) | 0 / 46 (0%) |  |
| # of positive axillary lymph nodes (n / N (%)) |  |  | 0.003<sup>2</sup> |  |  | 0.005<sup>2</sup> |
| 0 | 133 / 235 (57%) | 31 / 83 (37%) |  | 43 / 109 (39%) | 8 / 46 (17%) |  |
| 1 | 96 / 235 (41%) | 46 / 83 (55%) |  | 60 / 109 (55%) | 30 / 46 (65%) |  |
| 2 | 6 / 235 (2.6%) | 6 / 83 (7.2%) |  | 6 / 109 (5.5%) | 8 / 46 (17%) |  |
| Histologic TIL score (Median (IQR)) | 40 (20-60) | 30 (10-45) | 0.007<sup>1</sup> | 30 (15-45) | 23 (15-45) | 0.21<sup>1</sup> |
| Missing (n) | 1 | 1 |  | 0 | 2 |  |
| MHCII Immune Activation Score (Median (IQR)) | 1,501 (870-2,406) | 1,113 (579-1,826) | 0.005<sup>1</sup> | 1,517 (958-2,030) | 1,046 (738-1,746) | 0.022<sup>1</sup> |
| Missing (n) | 0 | 1 |  |  |  |  |

<sup>1</sup> Wilcoxon rank sum test
<sup>2</sup> Fisher's exact test

To further explore why TIL score and IA Score could have different prognostic power, we examined the correlations between these two immune biomarkers.

Histologic TIL score correlated positively with the IA Score in both subtypes (Figure 2, Basal-like r = 0.66, p = 1.5×10⁻⁴¹, HER2E r = 0.73, p = 2.4×10⁻²⁶). The strong correlation between IA Score and TILs indicates that the IA Score reflects the tumor immune microenvironment while substantial variability in IA at any given TIL level suggests IA Scores provide additional resolution beyond morphologic assessment alone. The dashed line at IA = 2400 highlights the previously determined cutpoint^33^ for identifying high IA, low risk patients. Many tumors with moderate TIL counts span both sides of this IA threshold, illustrating that the expression-based IA Score would provide different prognostic information than morphologic TIL assessment.

**Figure 2.**
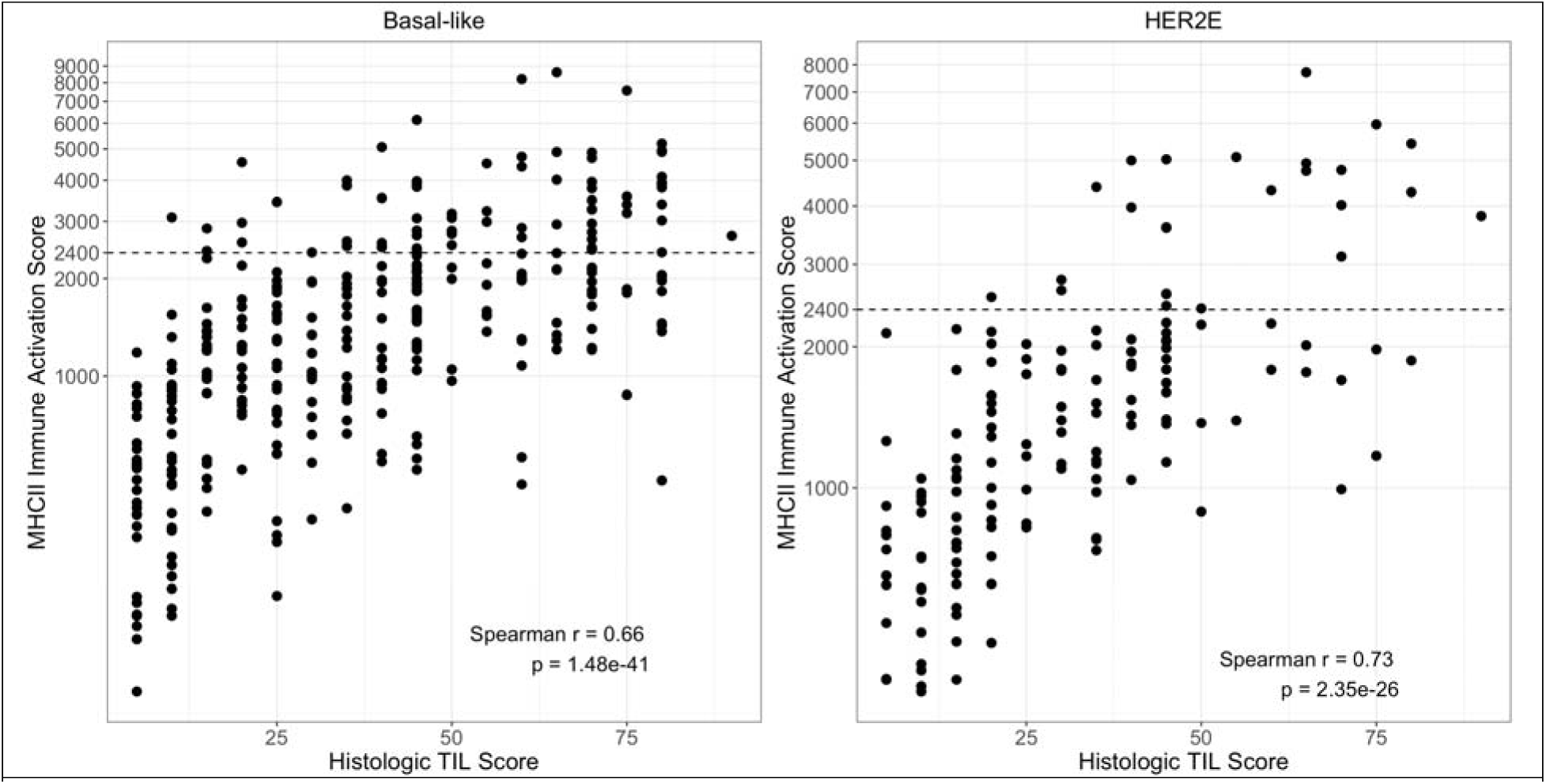
Correlation between IA Score (log10 scale) and Histologic TIL counting (linear scale) in Basal-like tumors (A, n=316) and HER2E tumors (B, n=153). Previously defined cutpoint (IA=2400) is horizontal dashed line. Spearman correlation coefficient and p-values are shown.

### MHCII-IA and TILs as Prognostic Variables

To determine which clinicopathologic variables were significant independent prognostic factors, multivariable Cox proportional hazards models accounting for age at diagnosis, lymph node positivity and either TIL scores>50% or log-transformed IA Score (log10[IA]) were computed for the Basal-like and HER2E groups separately (Figure 3). Analysis was restricted to patients for whom both histologic TIL score and IA Score were obtained. Age was not significantly associated with RFS in either subtype. Nodal positivity was consistently associated with worse RFS in both Basal-like and HER2E tumors (Basal-like: HR 2.04, 95% CI 1.31–3.19, p=0.002; HER2E: HR 2.71, 95% CI 1.26–5.84, p=0.011). In Basal-like tumors, high tumor-infiltrating lymphocytes (TILs ≥50%) were associated with improved RFS (HR 0.55, 95% CI 0.32–0.93, p=0.027); this association did not reach significance in HER2E tumors (HR 0.56, 95% CI 0.22–1.41, p=0.217). By comparison, higher IA Scores (log10[IA]) were associated with reduced risk of recurrence in Basal-like tumors (HR 0.45, 95% CI 0.25–0.83, p=0.010) and HER2E tumors (HR 0.24, 95% CI 0.08–0.69, p=0.008). These findings indicate nodal status and IA Score are significant independent prognostic factors in both Basal-like and HER2E breast cancer.

**Figure 3.**
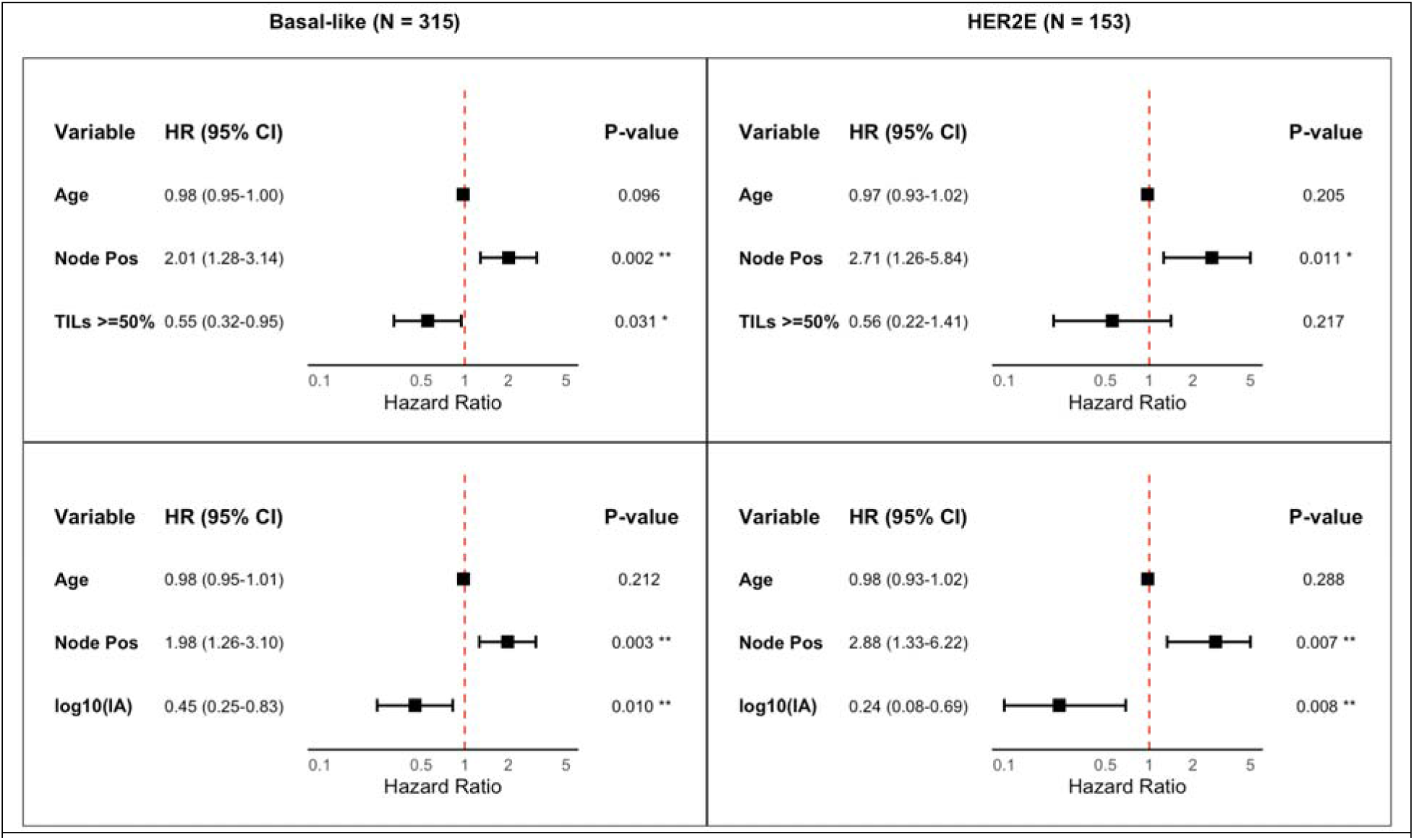
Multivariate Cox models of clinicopathologic variables and immune biomarkers with RFS. Forest plots show hazard ratios and 95% confidence intervals for Basal-like (left) and HER2E (right) tumors. Models included age, nodal status, and either TILs ≥50% (top) or log-transformed IA Score (bottom). Squares indicate hazard ratios, horizontal lines indicate 95% confidence intervals, and the dashed line denotes HR = 1. P-values are from Wald tests; *p < 0.05, **p < 0.01.

To evaluate the incremental prognostic value of immune biomarkers beyond standard clinicopathologic variables, we compared multivariable Cox models incorporating continuous TIL Score or IA Score within each intrinsic subtype (Table 2).

**Table 2.** Cox proportional hazards models were used to evaluate whether adding histologic tumor-infiltrating lymphocyte percentage (TIL Score) or IA Score (log10(IA Score)) variables improved prediction of recurrence-free survival beyond standard clinical factors (Age, T stage, and N stage) in patients for which both were measured (Basal-like N=315, HER2E N=153).

| Cox Model Comparison by Subtype |  |  |  |  |  |  |  |  |  |
| --- | --- | --- | --- | --- | --- | --- | --- | --- | --- |
| Model | Relative HR (95% CI) | ΔHR | C-index (95% CI) | ΔC-index | LR $\chi^2$ | LR p-value | AIC | Pseudo-$R^2$ | % Recur in Lowest Risk Quartile |
| Basal-like |  |  |  |  |  |  |  |  |  |
| Clinical (Age+T+N) | NA | NA | 0.643 (0.585-0.702) | NA | NA | NA | 890.995 | NA | 17.7 |
| Clinical + TIL Score | 0.985 (0.975-0.995) | -0.015 | 0.668 (0.61-0.726) | 0.024 | 8.510 | 0.004 | 884.485 | 0.027 | 15.2 |
| Clinical + log10(IA Score) | 0.469 (0.255-0.863) | -0.531 | 0.67 (0.613-0.727) | 0.026 | 5.697 | 0.017 | 887.297 | 0.018 | 11.4 |
| HER2E |  |  |  |  |  |  |  |  |  |
| Clinical (Age+T+N) | NA | NA | 0.656 (0.577-0.734) | NA | NA | NA | 419.056 | NA | 12.8 |
| Clinical + TIL Score | 0.986 (0.971-1.002) | -0.014 | 0.675 (0.599-0.751) | 0.019 | 3.217 | 0.073 | 417.839 | 0.021 | 12.8 |
| Clinical + log10(IA Score) | 0.192 (0.066-0.558) | -0.808 | 0.704 (0.63-0.777) | 0.048 | 9.477 | 0.002 | 411.579 | 0.060 | 10.3 |

In Basal-like tumors, inclusion of either the TIL Score or IA Score significantly improved model fit compared to the clinical model alone (LR χ² = 8.51, p = 0.004, and LR χ² = 5.70, p = 0.017, respectively). Both variables modestly increased model discrimination (C-index from 0.64 to 0.67) and explained variation (Pseudo-R² = 0.027–0.018). Notably, the percent of patients in the lowest risk quartile that recurred decreased from 17.7% in the clinical model to 15.2% with TIL Score and 11.4% with IA Score, highlighting improved identification of low-risk patients.

In HER2E tumors, adding the TIL Score did not significantly improve model performance (LR χ² = 3.22, p = 0.073), whereas including IA Score yielded a marked improvement (LR χ² = 9.48, p = 0.002), reducing Akaike Information Criterion (AIC) from 419.1 to 411.6 and increasing the C-index from 0.66 to 0.70. The percent of patients in the lowest risk quartile that recurred declined from 12.8% in the clinical model to 10.3% with IA Score, demonstrating enhanced stratification of low-risk patients.

To further examine how these biomarkers stratify risk across thresholds, we evaluated recurrence probability as a function of increasing TIL score and IA Score cutpoints, stratified by nodal status and subtype (Figure 4). Consistent with the Cox model results, increasing IA Score was associated with progressively lower recurrence risk in both Basal-like and HER2E tumors, with clear separation between node-negative and node-positive patients across the range of thresholds. In contrast, TILs demonstrated weaker and less consistent relationships with recurrence, particularly in HER2E disease, where risk estimates and confidence intervals were wide and variable across thresholds.

**Figure 4.**
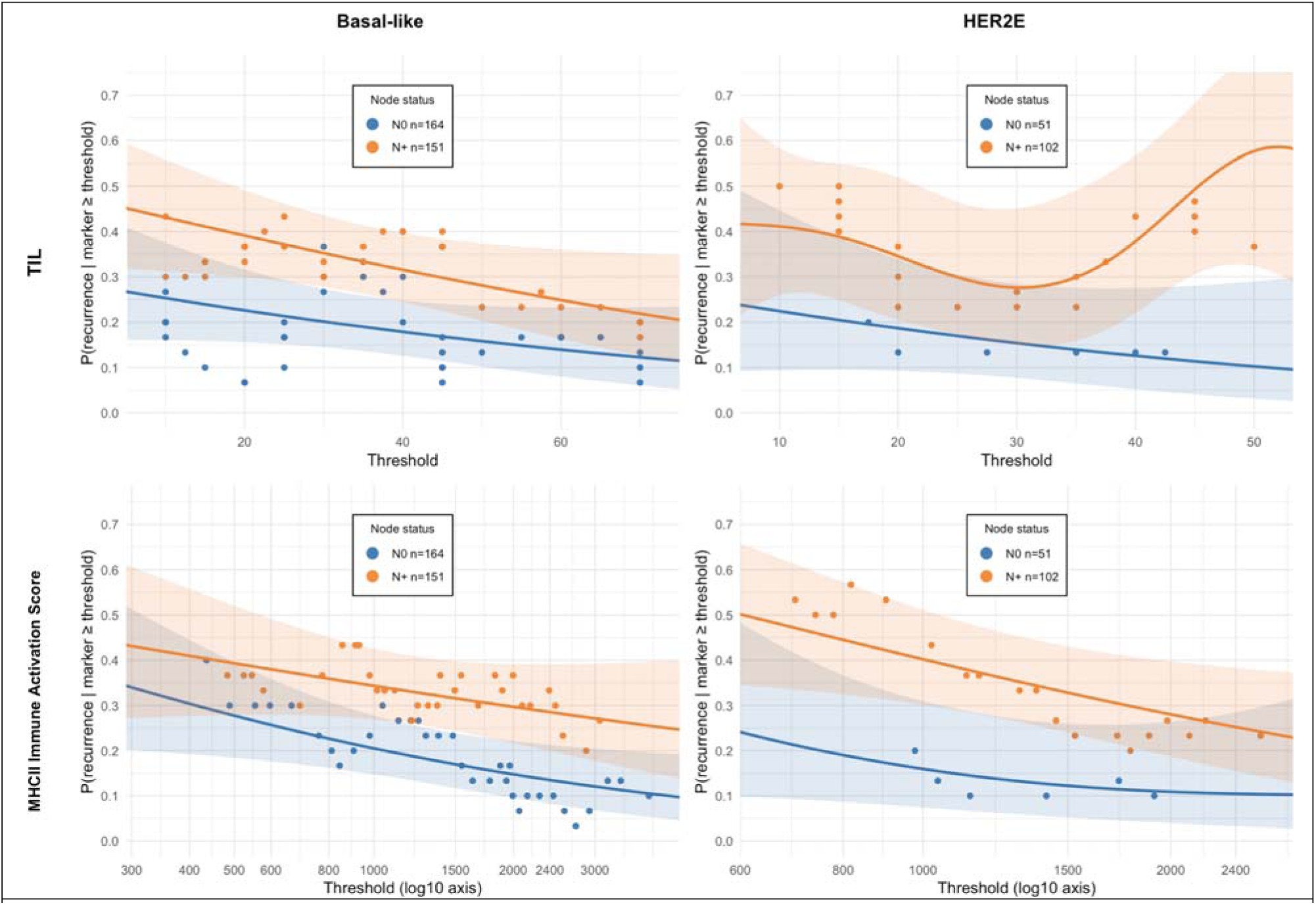
Cumulative probability of relapse is shown across increasing thresholds of histologic tumor-infiltrating lymphocytes (TIL; top row) and IA Score (bottom row). Points denote sliding-window estimates of observed recurrence risk, and shaded ribbons represent 95% bootstrap confidence intervals.

To examine the difference in RFS between patients with high and low IA Scores at specific threshold cutpoints, we used a cohort-wide median IA Score (50^th^ percentile IA=1370.905). The association with RFS was evaluated in each subtype and nodal status (Figure 5).

**Figure 5.**
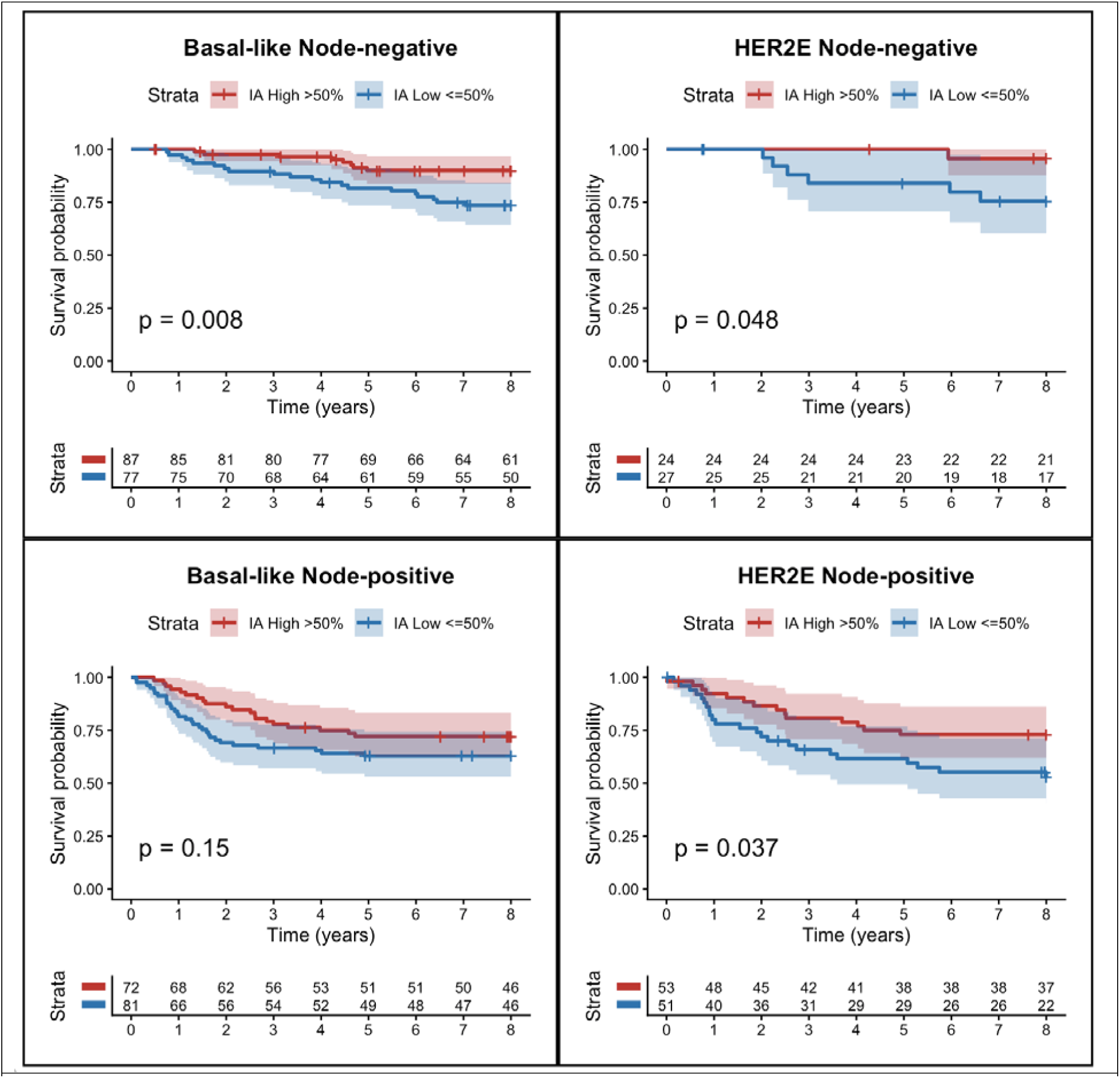
RFS stratified by median IA Score, subtype, and lymph node status. Kaplan–Meier curves show RFS censored at 8 years. Shaded areas represent 95% confidence intervals. P-values were calculated using the log-rank test. The number of patients at risk for each time interval is shown in the tables.

High IA Scores (>50th percentile) were associated with improved RFS across both subtypes and nodal groups, with statistically significant differences observed in node-negative Basal-like (p=0.008) and both node-negative and node-positive HER2E tumors (p=0.048 and p=0.037 respectively). Among node-negative patients, High IA Scores demonstrated particularly favorable outcomes, with ∼90% and ∼96% 8-year RFS in Basal-like and HER2E disease, respectively.

Notably, in node-positive disease, high IA Scores were associated with outcomes approaching those of node-negative patients. In the Basal-like group, the Node-Positive/High IA patients reached an 8-year RFS of 70.5%, approaching the level of the Node-Negative/Low IA subgroup (73.6%). Among patients with HER2E tumors, those with Node-Positive/High IA status achieved an 8-year RFS of 78.4%, numerically outperforming the Node-Negative/Low IA group (75.6%).

### MHCII-IA and Risk Assessment with Pre-specified Cutpoint

A previous study that applied the MHCII Immune Activation assay to institutional cohorts of Basal-like triple negative breast cancer patients defined a cutpoint for high IA Scores as greater than or equal to 2400 to identify patients who have a very low risk of recurrence^33^. To evaluate the clinical utility of that previously defined cutpoint for high IA Scores (IA>=2400) in these PAM50 classified Basal-like and HER2E tumors, RFS was examined for patients stratified on IA Score and lymph node status (Figure 6).

**Figure 6.**
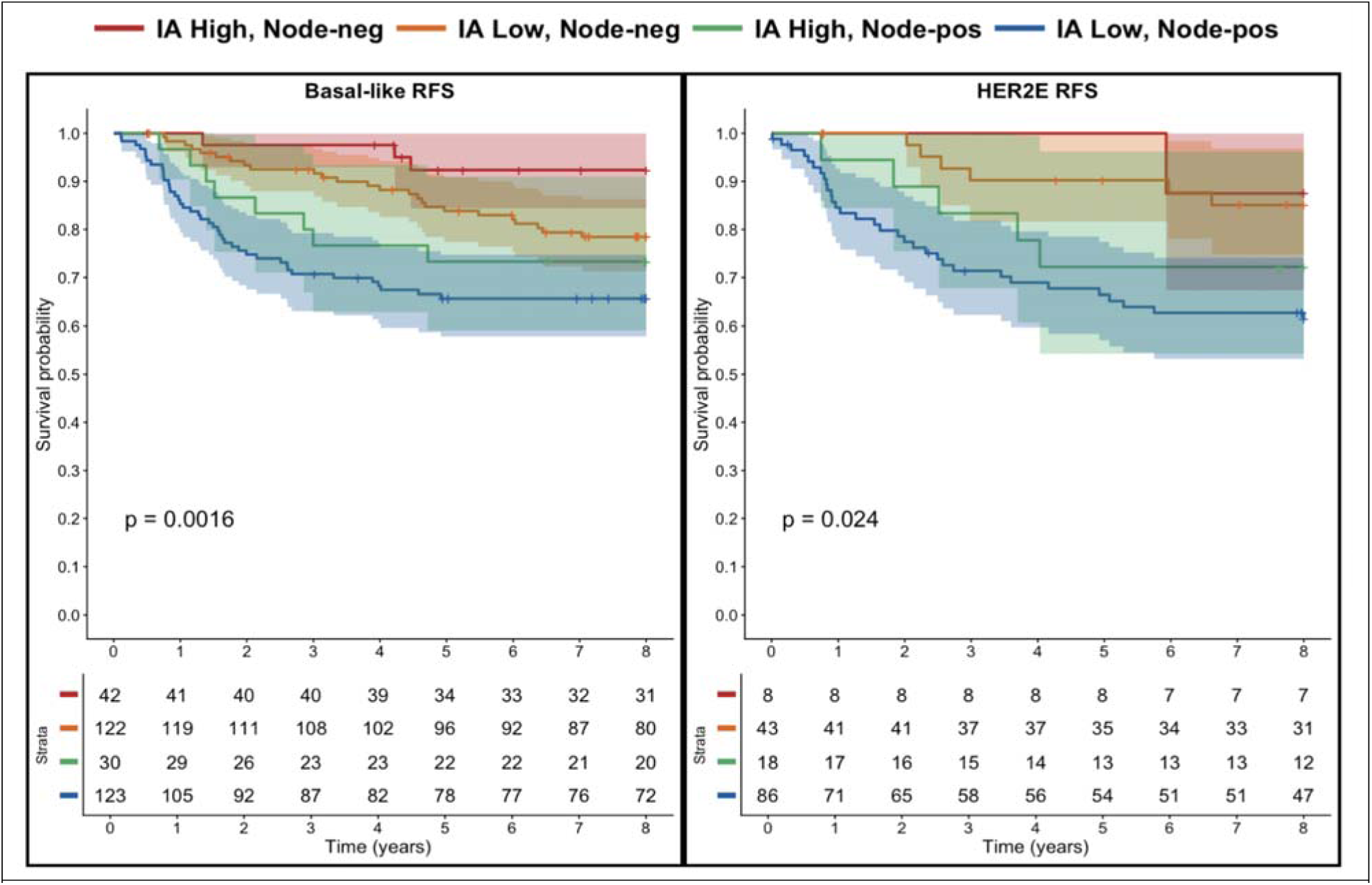
RFS outcomes using a cutpoint previously defined for high IA Scores in Basal-like breast tumors. Kaplan–Meier curves show 8-year RFS (top) in Basal-like (left) and HER2E (right) tumors, stratified by IA Score ≥2400 and lymph node status. Tick marks indicate censored observations, shaded areas show 95% confidence intervals, p-values are from log-rank tests, and risk tables show the number at risk over time.

High IA in the absence of nodal involvement (Red) identified a group with remarkable long-term outcomes; 92% 8-year RFS Basal-like and 88% 8-year RFS in HER2E. In contrast, the combination of a low IA Score and nodal involvement (Blue) identified a group at particularly high risk of recurrence with only 66% 8-year RFS in Basal-like and 62% 8-year RFS in HER2E.

High IA attenuated the adverse effect of nodal involvement in Basal-like tumors, where IA-high/node-positive patients (Green) had outcomes approaching IA-low/node-negative patients (Orange) with 73%-78% RFS at 8 years. This effect was less pronounced in HER2E disease at this higher cutpoint.

At this higher cutpoint, only 16.8% of HER2E patients and 22.7% of Basal-like patients are assigned into the high IA Score group. Interestingly, the rates of RFS for each subgroup are similar to the rates observed when the median value (IA=1370.905) was used as cutpoint in the exploratory analysis in Figure 5. This robust prognostic association is consistent with the continuous nature of IA Score depicted in Figure 4, and indicates that lower cutpoints could be considered, particularly for HER2E, if the goal is to assign the maximum number of low-risk patients into the high IA Score group.

Negative predictive value (NPV) is a crucial measure when evaluating a biomarker for therapy de-escalation. Using the predefined IA Score cutpoint of 2400, we observed consistently high NPV across subtypes (Table 3). The NPV for node-negative patients was 93% in Basal-like tumors and 88% in HER2E tumors. Importantly, high NPV was also maintained with the less stringent cohort-specific median IA threshold, with node-negative NPV of 91% and 96% for Basal-like and HER2E tumors, respectively. These findings demonstrate that the IA Score can reliably identify a subset of ER-negative breast cancer patients with a very low risk of recurrence, supporting its potential role in guiding safe de-escalation of chemotherapy.

**Table 3.**
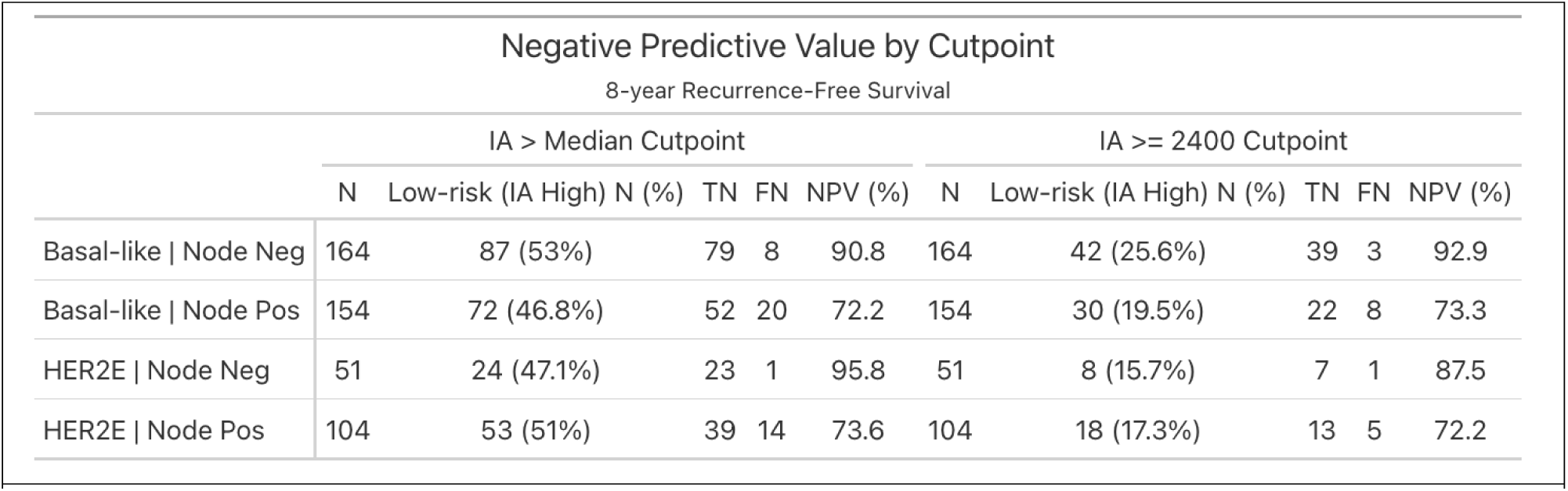
Negative Predictive Value and Size of the Low-Risk Population by IA Threshold (8-year RFS). Patients were classified as low risk based on IA Score thresholds (IA > median or IA ≥ 2400). True negatives (TN) denote patients in the low-risk group who did not recur. False negatives (FN) denote recurrences occurring in the low-risk group. Negative predictive value represents the proportion of patients without recurrence among those classified as low risk. The predefined IA ≥ 2400 threshold identifies a smaller, more stringently defined low-risk population compared to the median threshold.

### MHCII-IA and Taxane Prediction

Having established the prognostic value of the predefined IA Score threshold, we next sought to determine its association with benefit from adding paclitaxel to anthracycline-based chemotherapy. In MA21, the CEF and EC/T arms had superior RFS compared to the AC/T arm^35,37^. The MA21 AC/T arm was a less dose-intense, lower cumulative alkylator/anthracycline regimen compared to modern AC/T regimens. We chose to focus our analysis on the comparison of EC/T and CEF arms because previous analysis of the MA21 cohort used this same comparison to find that taxane benefit (EC/T vs CEF) was restricted to non-Luminal subtypes^37^.

We performed a subgroup analysis of 8-year RFS across three distinct cohorts defined by IA Scores and nodal involvement. Patients were stratified using the predefined IA cutpoint of 2400 and further by nodal status (Figure 7). The small number of patients who received adjuvant trastuzumab (shown in Table 1) were excluded from this analysis.

**Figure 7.**
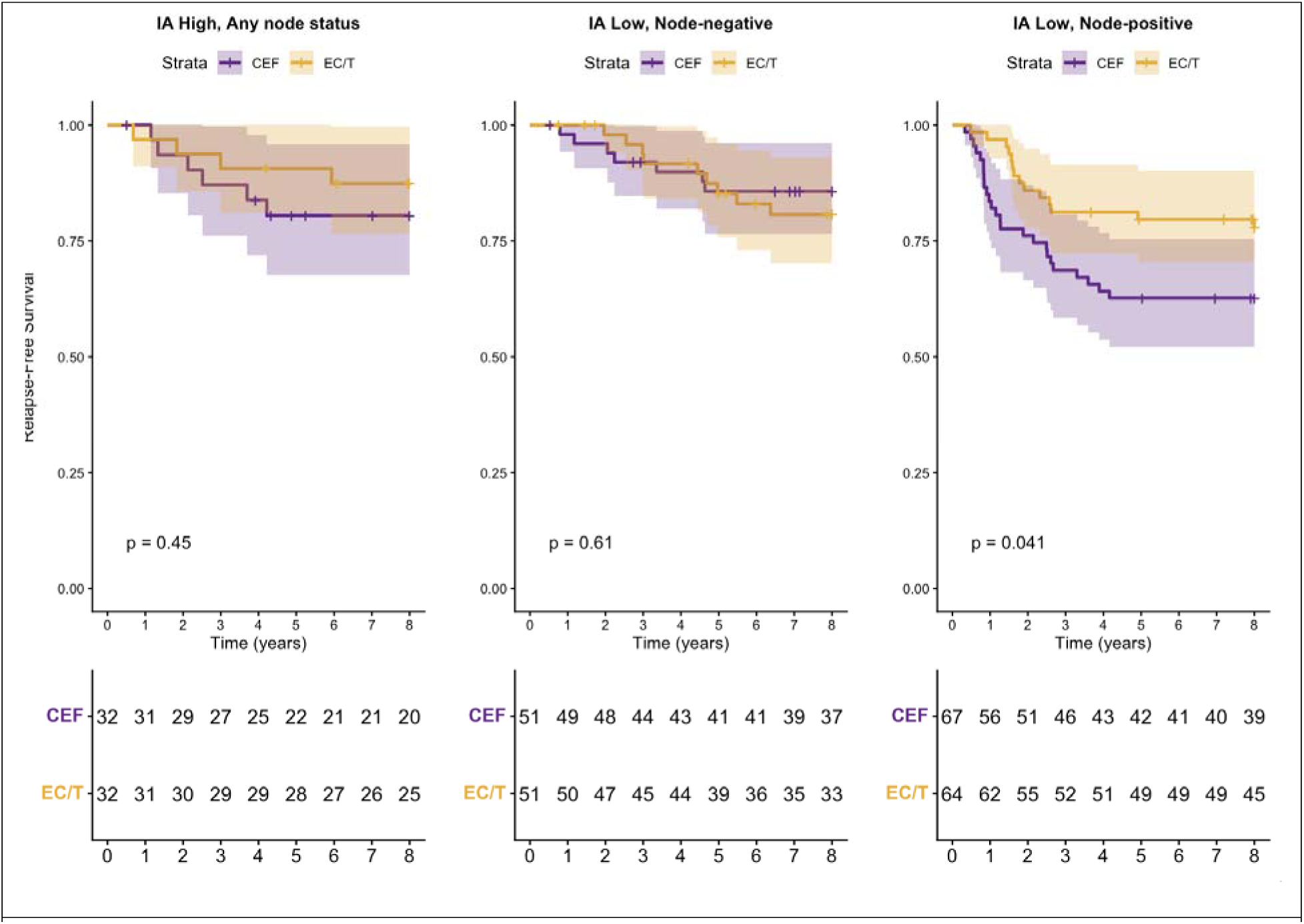
Association between adjuvant chemotherapy regimen and RFS stratified by IA Score and nodal status. Kaplan–Meier curves comparing patients treated with CEF (cyclophosphamide, epirubicin, and fluorouracil; purple) versus EC/T (epirubicin and cyclophosphamide followed by paclitaxel; gold). Patients were stratified using a predefined IA Score cutpoint of 2400, defining IA-High and IA-Low groups, and further subdivided by nodal status. Patients who received adjuvant trastuzumab were excluded from this analysis. Panels show IA-High (any nodal status) (left), IA-Low with node-negative disease (middle), and IA-Low with node-positive disease (right). Data were censored at 8 years of follow-up. Shaded areas represent 95% confidence intervals. P-values were calculated using the log-rank test. Numbers at risk are shown below each panel.

In patients with high IA Score (IA ≥ 2400), the addition of paclitaxel (EC/T) provided no incremental benefit over CEF regardless of nodal status with >80% 8-year RFS and no significant difference between treatment arms (log-rank p = 0.45; Figure 7, Left Panel). Similarly favorable outcomes (>80% RFS at 8 years) were observed for the Node-Negative low IA subgroup, with no significant difference between the treatment arms (log-rank p = 0.61, Figure 7, Middle Panel).

In contrast, a distinct benefit of adding paclitaxel was observed in the Node-Positive IA-Low cohort. In this high-risk, low IA Score group, patients receiving EC/T exhibited significantly improved RFS (78% at 8 years) compared to those receiving CEF (63% at 8 years) (log-rank p =0.041, Figure 7, Right Panel) with early and sustained separation of survival curves.

The observed treatment effect was confined to the node-positive, IA-low subgroup and was not observed among node-negative patients or among patients with high IA Scores. Because the clinical hypothesis involved both IA Score and nodal status, the relevant interaction extends beyond a simple treatment-by-IA model and requires consideration of treatment arm, IA group, and nodal status simultaneously. Formal interaction analyses were performed but were not statistically significant, likely reflecting the limited number of events within some strata (e.g., only 11 events in the IA-high group). We therefore focused on stratified analyses corresponding to the clinically defined groups shown in Figure 7. In stratified Cox models, a significant benefit from EC/T was observed only in the node-positive, IA-low subgroup (HR 0.52, 95% CI 0.28–0.94; p = 0.032). No treatment effect was observed among node-negative IA-low patients (HR 1.14, 95% CI 0.44–2.97; p = 0.783) or among patients with high IA Scores (HR 0.82, 95% CI 0.25–2.69; p = 0.742). These findings suggest that the previously reported taxane benefit in non-Luminal breast cancers may be concentrated within node-positive tumors with low IA Scores. IA high or node-negative patients have equally high rates of RFS regardless of whether they receive CEF or EC/T and derived no apparent benefit from the addition of paclitaxel to anthracycline chemotherapy.

## Discussion

The prognostic significance of high TILs and improved survival in the adjuvant setting after chemotherapy in early-stage TNBC has been well established^15^. In the neo-adjuvant setting, high TILs (≥60%) are associated with an increased probability of response to chemotherapy by ∼20% compared to low TILs in both ER-/HER2+ breast cancers and TNBC^40^. In this study, we demonstrate that the IA Score is a robust prognostic biomarker for identifying Basal-like and HER2E breast cancer patients at low risk of recurrence. The IA Score remained independently associated with recurrence risk after adjustment for clinicopathologic factors, and improved model discrimination when added to clinicopathologic variables in both subtypes. These findings validate prior reports of the prognostic significance of the IA Score in Basal-like breast cancer ^33^. This is also the first report of the prognostic significance of the IA Score in HER2E breast cancer, showing its relevance to HER2+ disease. When compared to histologic TIL score, the IA Score provided stronger and more consistent prognostic value, particularly in the HER2E subtype, likely reflecting its ability to capture both immune cell abundance and functional activation. IA Score is an immune biomarker with similar utility to TILs but circumvents the challenge of inter-observer variability and has improved prognostic power in HER2E breast cancer.

A particularly notable finding of this study is the high negative predictive value achieved by the IA Score in a population otherwise considered to be at high risk of recurrence. The ability to identify patients with a very low likelihood of relapse within high-risk subtypes such as Basal-like and HER2E disease is clinically significant, as it provides a potential framework for treatment de-escalation in settings where overtreatment remains common.

Notably, the MA.21 trial enrolled high-risk patients between 1997 and 2005 when chemotherapy was the only systemic anti-neoplastic therapy available for these breast cancer subtypes^35^. In this study we found that patients with high IA Scores and negative lymph nodes had excellent long-term outcomes despite being treated in a pre-trastuzumab and pre-pembrolizumab era, suggesting that strong endogenous immune activation is associated with favorable prognosis even in the absence of modern targeted therapies. Interestingly, the RFS rates achieved by the high IA Score subgroup are similar to those reported in contemporary trials incorporating HER2-targeted and immunotherapy-based regimens^41,42^, suggesting that these patients’ endogenous immune activation confers comparable clinical benefit. In node-positive disease, high IA Score was associated with improved outcomes relative to low IA Score tumors, with outcomes approaching those of node-negative low IA Score patients. This result suggests that endogenous immune activation may partially mitigate the risks associated with nodal involvement.

Beyond prognosis, IA Score was associated with differential benefit from taxane therapy, supporting its potential utility for risk stratification and treatment selection in these subtypes for which such biomarkers are currently lacking. Previous analysis of the MA21 cohort found that taxane benefit (EC/T vs CEF) was restricted to non-Luminal subtypes^37^, which are the focus of the current study, in which we found the benefit from taxane therapy was further enriched by IA Score and was only significant in patients with low IA Score node-positive disease. Patients with high IA Score and/or negative lymph nodes had equally high rates of RFS in the EC/T and CEF arms and derived no apparent benefit from the addition of paclitaxel to anthracycline chemotherapy.

Together, these results support the potential clinical utility of the IA Score for guiding treatment selection in Basal-like and HER2E breast cancer. Patients with high IA Score - particularly those with node-negative disease - are candidates for prospective trials evaluating de-escalation of taxanes and concomitant improvement in quality-of-life metrics. Across studies we have observed that 15-25% of Basal-like or HER2E breast cancer patients have high IA Scores and negative lymph nodes which we show is associated >90% RFS without paclitaxel, pembrolizumab, or trastuzumab in their treatment regimen. At the other end of the spectrum, patients with low IA Score and node-positive disease are at highest risk and benefit from taxanes. Randomized targeted therapy trials that selectively enroll these patients have the potential to demonstrate the greatest survival improvement.

This study has several limitations. While we applied a previously developed and locked down classifier onto materials from a randomized clinical trial, this is still a retrospective biomarker analysis, and treatment paradigms have evolved since MA.21 with the introduction of targeted therapies and immune checkpoint inhibitors. However, this context also enables assessment of the prognostic impact of endogenous immune activation independent of these therapies. Prospective studies in contemporary cohorts will be required to establish clinical utility in the context of modern regimens.

Now that pembrolizumab is standard of care for TNBC^42^ and the long-term burden of immune-related adverse events (irAEs) is mounting in oncology and rheumatology clinics, the utility of IA Score for immunotherapy selection is an important future direction^43^. Testing for immune biomarkers (e.g. PDL1) in metastatic TNBC has been shown to be important for predicting response to immuno-blockade therapy^22^ but it is not clear if some PDL1+ patients would have similarly favorable outcomes without the immunotherapy. Specifically, can IA Score distinguish patients who have endogenous immune activation and an inherently good prognosis who can safely avoid the risk of irAEs from patients who require the addition of neoadjuvant pembrolizumab to achieve anti-tumor immune activation and long-term RFS?

Mechanistic and functional studies using mouse models of cancer provide evidence for how MHCII expression in tumor cells supports endogenous anti-tumor immunity and long-term RFS. When tumor cells are engineered to express MHCII and implanted into syngeneic mice, they recruit CD4+ T cells and CD8+ T cells, induce a Th1 anti-tumor immune response, and inhibit disease progression without the need for chemotherapy or targeted therapy^28-32,44^. Interferon gamma, usually secreted by T cells, induces expression of the MHCII pathway in breast cancer cells^24^. Administering interferon gamma to mouse models of breast and ovarian cancer stimulated anti-tumor immunity and reduced the rate of tumor recurrence^45^. Together these functional studies support the concept that MHCII expression on tumor cells is both a marker of, and a contributor to, productive anti-tumor immunity. These data also pose an intriguing question: could careful dosing of neoadjuvant interferon gamma increase the number of patients with high IA Scores and long-term RFS?

Future studies are warranted to determine if the IA Score is prognostic and predictive in other cancer types where MHCII expression has been associated with good outcomes including melanoma^34^, ovarian cancer^46^, non-small cell lung cancer^47^, colorectal cancer^48,49^, and bladder cancer^50^.

## Acknowledgements

This work was financially supported by the National Cancer Institute of the National Institutes of Health under Award Number UH3CA262221. Thanks to Angela Snow, Austin Wood, and Sean Tavtigian in the Huntsman Cancer Institute for assisting with use of their liquid handling robot to aliquot the RNA specimens. Thanks to Anca Franzini in the Huntsman Cancer Institute for aliquoting the positive control RNA and shipping the plates. Thanks to Vijay Baichwal at Canopy Biosciences - A Bruker Company for running the NanoString instruments. Generative AI was used to review and revise the manuscript text for clarity.

## Author Contributions

PSB, LES, TON, and KEV designed the study. DG performed the histologic TIL assessment. BEC and KEV prepared and analyzed the data. PSB, TON, KEV drafted and edited the manuscript.

## References

1 Nielsen, T. O. et al. A comparison of PAM50 intrinsic subtyping with immunohistochemistry and clinical prognostic factors in tamoxifen-treated estrogen receptor-positive breast cancer. Clin Cancer Res 16, 5222–5232, doi:10.1158/1078-0432.CCR-10-1282 (2010).

2 Paik, S. et al. A multigene assay to predict recurrence of tamoxifen-treated, node-negative breast cancer. N Engl J Med 351, 2817–2826, doi:10.1056/NEJMoa041588 (2004).

3 Cardoso, F. et al. 70-Gene Signature as an Aid to Treatment Decisions in Early-Stage Breast Cancer. N Engl J Med 375, 717–729, doi:10.1056/NEJMoa1602253 (2016).

4 Goetz, M. P. et al. A two-gene expression ratio of homeobox 13 and interleukin-17B receptor for prediction of recurrence and survival in women receiving adjuvant tamoxifen. Clin Cancer Res 12, 2080–2087, doi:10.1158/1078-0432.CCR-05-1263 (2006).

5 Nielsen, T. et al. Analytical validation of the PAM50-based Prosigna Breast Cancer Prognostic Gene Signature Assay and nCounter Analysis System using formalin-fixed paraffin-embedded breast tumor specimens. BMC Cancer 14, 177, doi:10.1186/1471-2407-14-177 (2014).

6 Dubsky, P. et al. The EndoPredict score provides prognostic information on late distant metastases in ER+/HER2- breast cancer patients. Br J Cancer 109, 2959–2964, doi:10.1038/bjc.2013.671 (2013).

7 Bottosso, M. et al. Gene Expression Assays to Tailor Adjuvant Endocrine Therapy for HR+/HER2- Breast Cancer. Clin Cancer Res 30, 2884–2894, doi:10.1158/1078-0432.CCR-23-4020 (2024).

8 Andre, F. et al. Biomarkers for Adjuvant Endocrine and Chemotherapy in Early-Stage Breast Cancer: ASCO Guideline Update. J Clin Oncol 40, 1816–1837, doi:10.1200/JCO.22.00069 (2022).

9 Henderson, I. C. et al. Improved outcomes from adding sequential Paclitaxel but not from escalating Doxorubicin dose in an adjuvant chemotherapy regimen for patients with node-positive primary breast cancer. J Clin Oncol 21, 976–983, doi:10.1200/JCO.2003.02.063 (2003).

10 Martin, M. et al. Randomized phase 3 trial of fluorouracil, epirubicin, and cyclophosphamide alone or followed by Paclitaxel for early breast cancer. J Natl Cancer Inst 100, 805–814, doi:10.1093/jnci/djn151 (2008).

11 Mackey, J. R. et al. Adjuvant docetaxel, doxorubicin, and cyclophosphamide in node-positive breast cancer: 10-year follow-up of the phase 3 randomised BCIRG 001 trial. Lancet Oncol 14, 72–80, doi:10.1016/S1470-2045(12)70525-9 (2013).

12. Early Breast Cancer Trialists’ Collaborative Group. Electronic address, b. o. c. o. a. u. & Early Breast Cancer Trialists’ Collaborative, G. Anthracycline-containing and taxane-containing chemotherapy for early-stage operable breast cancer: a patient-level meta-analysis of 100 000 women from 86 randomised trials. Lancet 401, 1277-1292, doi:10.1016/S0140-6736(23)00285-4 (2023).

13 Winters-Stone, K. M. et al. Falls, Functioning, and Disability Among Women With Persistent Symptoms of Chemotherapy-Induced Peripheral Neuropathy. J Clin Oncol 35, 2604–2612, doi:10.1200/JCO.2016.71.3552 (2017).

14 Seretny, M. et al. Incidence, prevalence, and predictors of chemotherapy-induced peripheral neuropathy: A systematic review and meta-analysis. Pain 155, 2461–2470, doi:10.1016/j.pain.2014.09.020 (2014).

15 Loi, S. et al. Tumor-Infiltrating Lymphocytes and Prognosis: A Pooled Individual Patient Analysis of Early-Stage Triple-Negative Breast Cancers. J Clin Oncol 37, 559–569, doi:10.1200/JCO.18.01010 (2019).

16 Schlam, I., Loi, S., Salgado, R. & Swain, S. M. Tumor-infiltrating lymphocytes in HER2-positive breast cancer: potential impact and challenges. ESMO Open 10, 104120, doi:10.1016/j.esmoop.2024.104120 (2025).

17 Salgado, R. et al. The evaluation of tumor-infiltrating lymphocytes (TILs) in breast cancer: recommendations by an International TILs Working Group 2014. Ann Oncol 26, 259–271, doi:10.1093/annonc/mdu450 (2015).

18 de Jong, V. M. T. et al. Prognostic Value of Stromal Tumor-Infiltrating Lymphocytes in Young, Node-Negative, Triple-Negative Breast Cancer Patients Who Did Not Receive (neo)Adjuvant Systemic Therapy. J Clin Oncol 40, 2361–2374, doi:10.1200/JCO.21.01536 (2022).

19 El Bairi, K., et al. The tale of TILs in breast cancer: A report from The International Immuno-Oncology Biomarker Working Group. NPJ Breast Cancer 7, 150, doi:10.1038/s41523-021-00346-1 (2021).

20 Leon-Ferre, R. A. et al. Tumor-Infiltrating Lymphocytes in Triple-Negative Breast Cancer. JAMA 331, 1135–1144, doi:10.1001/jama.2024.3056 (2024).

21 Rizzo, A. et al. KEYNOTE-522, IMpassion031 and GeparNUEVO: changing the paradigm of neoadjuvant immune checkpoint inhibitors in early triple-negative breast cancer. Future Oncol 18, 2301-2309, doi:10.2217/fon-2021-1647 (2022).

22 Cortes, J. et al. Pembrolizumab plus chemotherapy versus placebo plus chemotherapy for previously untreated locally recurrent inoperable or metastatic triple-negative breast cancer (KEYNOTE-355): a randomised, placebo-controlled, double-blind, phase 3 clinical trial. Lancet 396, 1817–1828, doi:10.1016/S0140-6736(20)32531-9 (2020).

23 Schmid, P. et al. Pembrolizumab plus chemotherapy as neoadjuvant treatment of high-risk, early-stage triple-negative breast cancer: results from the phase 1b open-label, multicohort KEYNOTE-173 study. Ann Oncol 31, 569–581, doi:10.1016/j.annonc.2020.01.072 (2020).

24 Forero, A. et al. Expression of the MHC Class II Pathway in Triple-Negative Breast Cancer Tumor Cells Is Associated with a Good Prognosis and Infiltrating Lymphocytes. Cancer Immunol Res 4, 390–399, doi:10.1158/2326-6066.CIR-15-0243 (2016).

25 Park, I. A. et al. Expression of the MHC class II in triple-negative breast cancer is associated with tumor-infiltrating lymphocytes and interferon signaling. PLoS One 12, e0182786, doi:10.1371/journal.pone.0182786 (2017).

26 Stewart, R. L., Matynia, A. P., Factor, R. E. & Varley, K. E. Spatially-resolved quantification of proteins in triple negative breast cancers reveals differences in the immune microenvironment associated with prognosis. Sci Rep 10, 6598, doi:10.1038/s41598-020-63539-x (2020).

27 Frangione, V., Mortara, L., Castellani, P., De Lerma Barbaro, A. & Accolla, R. S. CIITA-driven MHC-II positive tumor cells: preventive vaccines and superior generators of antitumor CD4+ T lymphocytes for immunotherapy. Int J Cancer 127, 1614–1624, doi:10.1002/ijc.25183 (2010).

28 Meazza, R., Comes, A., Orengo, A. M., Ferrini, S. & Accolla, R. S. Tumor rejection by gene transfer of the MHC class II transactivator in murine mammary adenocarcinoma cells. Eur J Immunol 33, 1183–1192, doi:10.1002/eji.200323712 (2003).

29 Mortara, L. et al. CIITA-induced MHC class II expression in mammary adenocarcinoma leads to a Th1 polarization of the tumor microenvironment, tumor rejection, and specific antitumor memory. Clin Cancer Res 12, 3435–3443, doi:10.1158/1078-0432.CCR-06-0165 (2006).

30 Armstrong, T. D., Clements, V. K. & Ostrand-Rosenberg, S. MHC class II-transfected tumor cells directly present antigen to tumor-specific CD4+ T lymphocytes. J Immunol 160, 661–666 (1998).

31 Pulaski, B. A. & Ostrand-Rosenberg, S. Reduction of established spontaneous mammary carcinoma metastases following immunotherapy with major histocompatibility complex class II and B7.1 cell-based tumor vaccines. Cancer Res 58, 1486–1493 (1998).

32 Thompson, J. A. et al. Tumor cells transduced with the MHC class II Transactivator and CD80 activate tumor-specific CD4+ T cells whether or not they are silenced for invariant chain. Cancer Res 66, 1147–1154, doi:10.1158/0008-5472.CAN-05-2289 (2006).

33 Stewart, R. L. et al. A Multigene Assay Determines Risk of Recurrence in Patients with Triple-Negative Breast Cancer. Cancer Res 79, 3466–3478, doi:10.1158/0008-5472.CAN-18-3014 (2019).

34 Johnson, D. B. et al. Melanoma-specific MHC-II expression represents a tumour-autonomous phenotype and predicts response to anti-PD-1/PD-L1 therapy. Nat Commun 7, 10582, doi:10.1038/ncomms10582 (2016).

35 Burnell, M. et al. Cyclophosphamide, epirubicin, and Fluorouracil versus dose-dense epirubicin and cyclophosphamide followed by Paclitaxel versus Doxorubicin and cyclophosphamide followed by Paclitaxel in node-positive or high-risk node-negative breast cancer. J Clin Oncol 28, 77–82, doi:10.1200/JCO.2009.22.1077 (2010).

36 Parker, J. S. et al. Supervised risk predictor of breast cancer based on intrinsic subtypes. J Clin Oncol 27, 1160–1167, doi:10.1200/JCO.2008.18.1370 (2009).

37 Liu, S. et al. Prognostic and predictive investigation of PAM50 intrinsic subtypes in the NCIC CTG MA.21 phase III chemotherapy trial. Breast Cancer Res Treat 149, 439-448, doi:10.1007/s10549-014-3259-1 (2015).

38 Liu, S. et al. Role of Cytotoxic Tumor-Infiltrating Lymphocytes in Predicting Outcomes in Metastatic HER2-Positive Breast Cancer: A Secondary Analysis of a Randomized Clinical Trial. JAMA Oncol 3, e172085, doi:10.1001/jamaoncol.2017.2085 (2017).

39 Riaz, N. et al. Prognostic and predictive capacity of tumor infiltrating lymphocytes in the MA.20 regional node radiotherapy trial. NPJ Breast Cancer 11, 97, doi:10.1038/s41523-025-00821-z (2025).

40 Denkert, C. et al. Tumour-infiltrating lymphocytes and prognosis in different subtypes of breast cancer: a pooled analysis of 3771 patients treated with neoadjuvant therapy. Lancet Oncol 19, 40–50, doi:10.1016/S1470-2045(17)30904-X (2018).

41 Cameron, D. et al. 11 years’ follow-up of trastuzumab after adjuvant chemotherapy in HER2-positive early breast cancer: final analysis of the HERceptin Adjuvant (HERA) trial. Lancet 389, 1195–1205, doi:10.1016/S0140-6736(16)32616-2 (2017).

42 Schmid, P. et al. Overall Survival with Pembrolizumab in Early-Stage Triple-Negative Breast Cancer. N Engl J Med 391, 1981–1991, doi:10.1056/NEJMoa2409932 (2024).

43 Kok, M. et al. Academic Uphill Battle to Personalize Treatment for Patients With Stage II/III Triple-Negative Breast Cancer. J Clin Oncol 42, 3523–3529, doi:10.1200/JCO.24.00372 (2024).

44 Ilkovitch, D. & Ostrand-Rosenberg, S. MHC class II and CD80 tumor cell-based vaccines are potent activators of type 1 CD4+ T lymphocytes provided they do not coexpress invariant chain. Cancer Immunol Immunother 53, 525–532, doi:10.1007/s00262-003-0486-4 (2004).

45 Sun, L. et al. Activating a collaborative innate-adaptive immune response to control metastasis. Cancer Cell 39, 1361–1374 e1369, doi:10.1016/j.ccell.2021.08.005 (2021).

46 Perez-Villatoro, F. et al. Single-Cell Spatial Atlas of High-Grade Serous Ovarian Cancer Uncovers MHC Class II as a Key Predictor of Spatial Tumor Ecosystems and Clinical Outcomes. Cancer Discov 16, 1100–1125, doi:10.1158/2159-8290.CD-25-1492 (2026).

47 Johnson, A. M. et al. Cancer Cell-Specific Major Histocompatibility Complex II Expression as a Determinant of the Immune Infiltrate Organization and Function in the NSCLC Tumor Microenvironment. J Thorac Oncol 16, 1694–1704, doi:10.1016/j.jtho.2021.05.004 (2021).

48 Matsushita, K. et al. Strong HLA-DR antigen expression on cancer cells relates to better prognosis of colorectal cancer patients: Possible involvement of c-myc suppression by interferon-gamma in situ. Cancer Sci 97, 57–63, doi:10.1111/j.1349-7006.2006.00137.x (2006).

49 Sconocchia, G. et al. HLA class II antigen expression in colorectal carcinoma tumors as a favorable prognostic marker. Neoplasia 16, 31–42, doi:10.1593/neo.131568 (2014).

50 Yi, R. et al. MHC-II Signature Correlates With Anti-Tumor Immunity and Predicts anti-PD-L1 Response of Bladder Cancer. Front Cell Dev Biol 10, 757137, doi:10.3389/fcell.2022.757137 (2022).

